# The COVID-19 Pandemic Impact on Primary Health Care services: An Experience from Qatar

**DOI:** 10.1101/2020.07.22.20160333

**Authors:** Mariam Abdulmalik, Mohamed Ghaith Al Kuwari, Samya Al Abdulla, Ahmad Haj Bakri, John Gibb, Mujeeb Chettiyam Kandy

## Abstract

**Introduction:** In March 2020, Qatar started reporting increased numbers of COVID-19 positive cases. The Ministry of Public Health in Qatar has developed an emergency action plan to respond to the outbreak with the Primary Health Care Corporation (PHCC) as a main component of that response.

**Aim:** The aim of this review is to understand and document the Impact of COVID 19 on PHCC in Qatar in terms of response, modifications of services and introduction of new alternatives

**Methodology:** A retrospective data analysis was conducted for all the COVID-19 swabbing activities and for all the utilization services volume across the PHCC health centers between January 2018 and May 2020.

**Results:** PHCC allocated testing sites for COVID-19 resulted in conducting 54824 swabs with 11455 positive cases and positivity rate of 20.8% between 14th of March and 15th of June 2020. The overall PHCC services utilization declined with overall reduction of 50% in April 2020. Alternative virtual and remote services were provided, telemedicine was introduced, and it made up 50% of the consultation volumes for April 2020. Home refill delivery medications managed to provide a total of 20920 delivered prescriptions by end of May 2020.

**Conclusion and recommendations:** To decrease the risk of infection to the patients and health care workers, PHCC in Qatar cancelled the appointments for some high-risk population. However, PHCC introduced virtual remote services that managed to make up for the in-person utilization volume and reflected acceptance in patients’ behaviours. PHCC continued in detecting positive COVID-19 cases among its targeted communities.

## Introduction

Coronaviruses are RNA viruses that are found in human beings and other mammals^1^. Even though most human coronavirus infections result in mild diseases, the world has witnessed two major epidemics in the past two decades from two different betacoronaviruses; severe acute respiratory syndrome coronavirus (SARS-CoV) and Middle East respiratory syndrome coronavirus (MERS-CoV). The latter two outbreaks collectively resulted in more than 10000 cases, with fatality rate of 10% and 37% for SARS-CoV and MERS-CoV, respectively^2^.

In December 2019, China reported to the WHO cases of unknown cause of pneumonia occurring in Wuhan, Hubei^3^. The samples viral genetic sequencing indicated a novel coronavirus^4^. The novel virus was named 2019 novel coronavirus (COVID-19) and 75-80% resemblance to SARS-CoV was confirmed^2^. As of June 9, 2020, approximately 7 million cases and over 400,000 deaths have been reported globally^5^. The COVID-19 outbreak was declared by the WHO as a public health emergency of international concern and the WHO put in place a series of temporary recommendations^6^. With no availability for specific antiviral therapy, efforts continue to develop antivirals and vaccine. Early indications suggest that the primary reservoir for the virus are the bats, given the close similarity to bat coronavirus^7^, while the efforts to identify the zoonotic of the virus continue, the public health measures for managing the outbreak rely on the existing preparedness national and regional capacities to prevent, detect, and response^8^.

Countries have been enhancing preparedness through the implementation and regular assessment of their national capacities to mitigate the effect of public health emergencies, including the emergence of a novel pathogen^9 10^.

The increased understanding of the capacities that countries have to prevent, detect, and respond to public health events has been possible through applying a monitoring and evaluation framework and ensuring that regular reporting of the response is in place^11^.

## Primary Care and COVID-19 pandemic in Qatar

In March 2020, Qatar started reporting increased numbers of COVID-19 positive cases. At that stage national restrictions were put in place. The Ministry of Public Health in Qatar has developed an emergency action plan to respond to the outbreak of COVID-19 with the Primary Health Care Corporation (PHCC) as a main component of that response. As of June 17, 2020, a total of 83,092 cases and 82 deaths have been reported in Qatar^12^.

Primary Health Care Corporation (PHCC), the main primary care provider in Qatar is serving 1.4 million individuals throughout a network of 27 primary health care centers covering all three main regions in the country. The services range from preventive services e.g. cancer screening, immunization, lifestyle counselling to therapeutic services for long term conditions, antenatal, and urgent care for adults and children. In addition to that PHCC provides general dental services, pharmacy, and laboratory services.

PHCC responded rapidly to the pandemic by opening the first COVID-19 centre for testing and holding on the 12^th^ of March 2020; and suspending all non-essential services and maintained only urgent services and walk in clinics as operational. Laboratory, pharmacy, diagnostics were all operational to support walk ins patients. As the epidemic continued, PHCC started to open more centers for testing and initiated new alternative services to respond to the needs of its target population.

The COVID-19 pandemic has heavily impacted how primary health care services have been delivered. This impact has positive and negative sides for the services and patients. To decrease the risk of infection to the patients and health care workers, Primary Health Care Corporation (PHCC) in Qatar had to cancel the booked appointments for some high-risk population e.g. NCD and antenatal visits, and the preventive visits e.g. screening and wellness. At the same time PHCC replaced the inperson consultations with telemedicine consultations. The pandemic affected the patients’ behaviour due to either adherence to the recommendations to stay at home, or their perceived risk of infection.

## Aim

The aim of this review is to understand and document the Impact of COVID 19 on Primary Health Care Corporation (PHCC) in Qatar in terms of PHCC response, modifications of services and clinical pathways, and the introduction so new alterative services.

### Methodology

A retrospective data analysis was conducted for all the COVID-19 swabbing activities and for all the utilization services volume across the 27 PHCC health centers to monitor the PHCC response to COVID-19 and to track the service utilization during the pandemic. The data was extracted from the business health intelligence between January 2018 and May 2020. Positivity rates among the screened patients and utilization trends were established to track the differences prior and post COVID-19 outbreak.

## Results

### 1. Primary health care Corporation response to COVID-19

In the beginning of March 2020 when the COVID-19 cases started rising in Qatar, the PHCC on the 12th of March 2020, designated Muaither health center as an exclusive COVID-19 testing and holding center. Then subsequently, it designated Rawdat Al Khail, Um Slal and Gharaffat al Rayan as COVID-19 testing and holding centers as of 18th of March and 9th of April 2020, respectively. In addition, one testing room at least was allocated within all other remaining health centers. These facilities helped in testing 54,824 individuals and detecting 11,455 confirmed cases, with highest positivity rate in May (see figure No.1 and No.2).

**Figure No.1:**
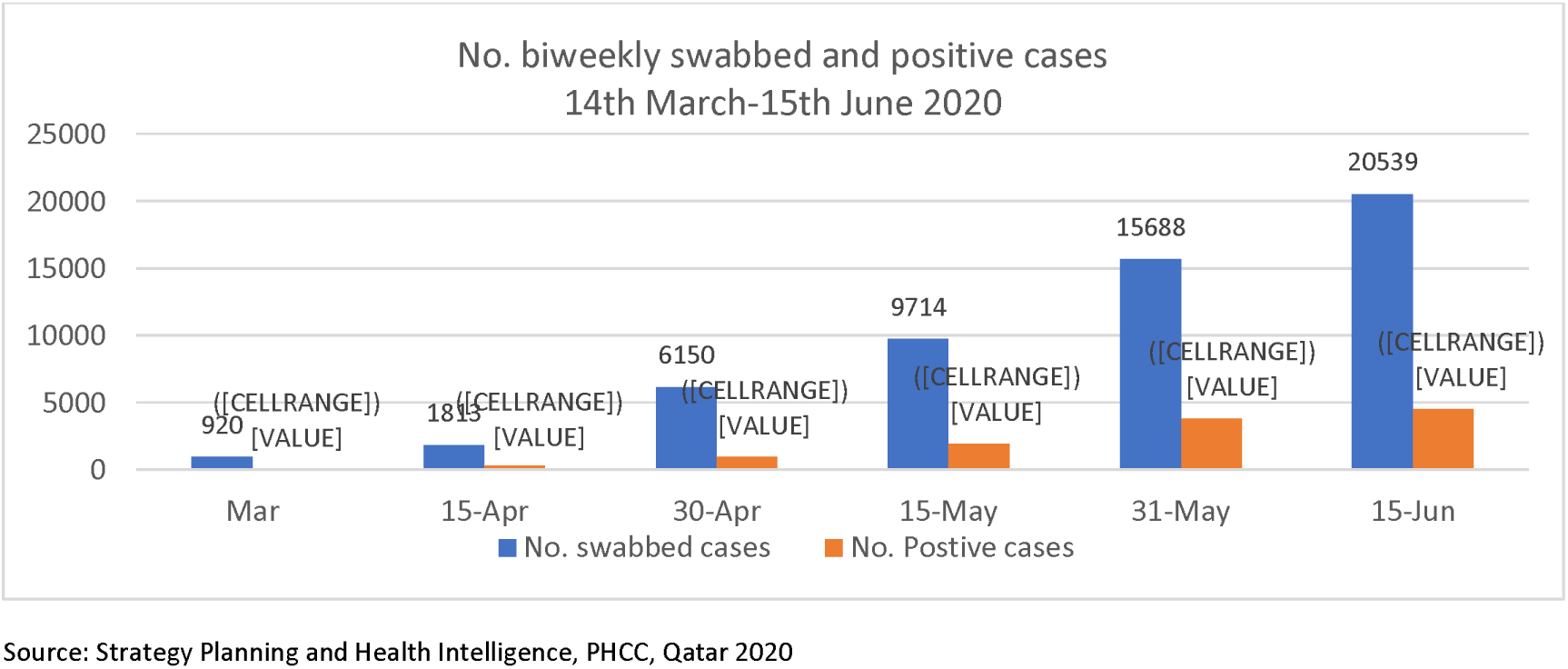
Number of biweekly PHCC swabbed and positive cases between the 14^th^ of March and the 15^th^ of June 2020.

**Figure No.2:**
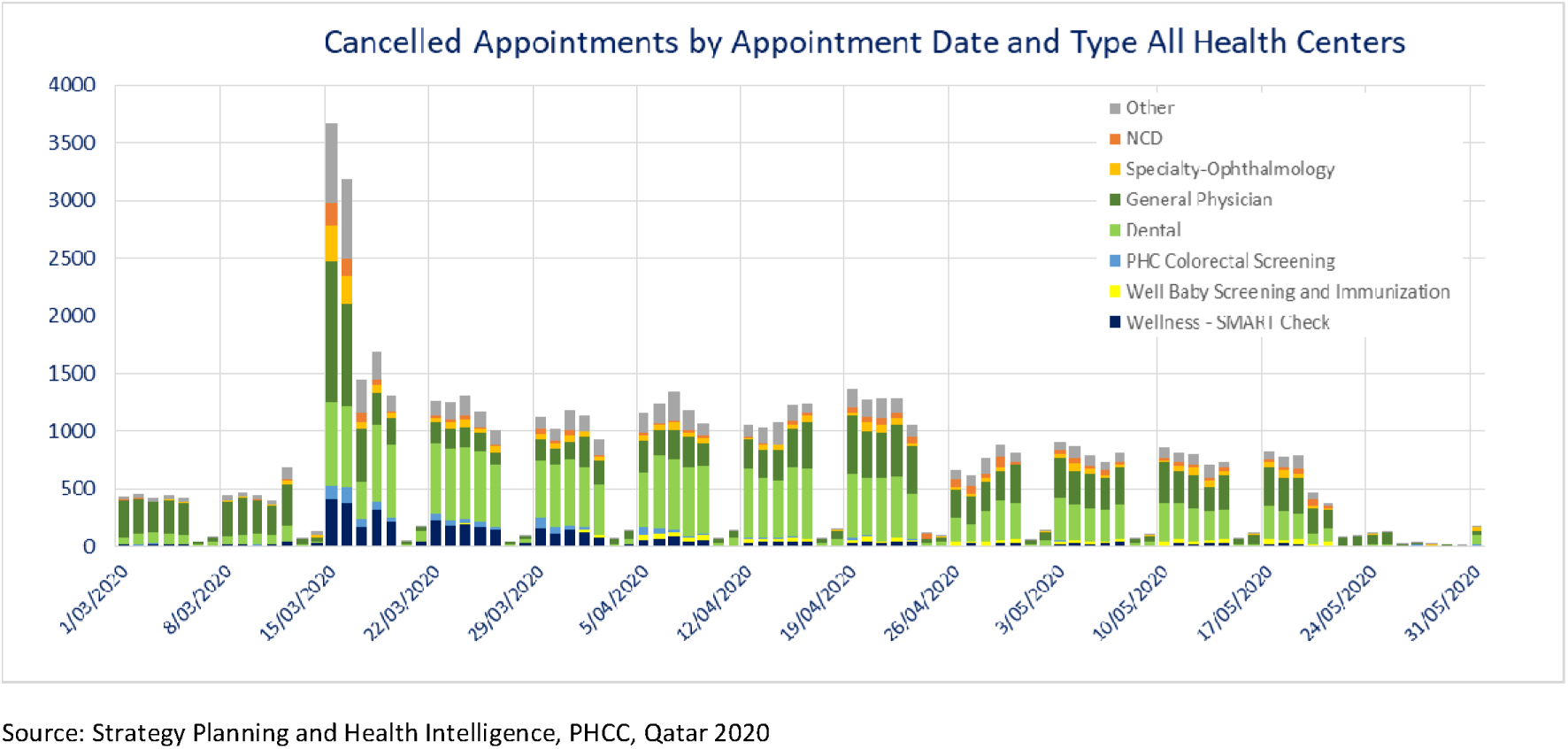
Cancelled appointments by appointment date and service type between 1^st^ of March and 30^th^ May 2020 for all health centers.

### 2. PHCC clinical pathways modification of services due to COVID-19

In mid-March 2020, PHCC cancelled all non-urgent appointments. Only the urgent services and walk-in clinics remained operational with the laboratory, pharmacy and all diagnostics being operational to support the walk-in services. The latter resulted in cancelling a total of 77,927 scheduled appointments by the end of April 2020 (see figure No.3).

**Figure No.3:**
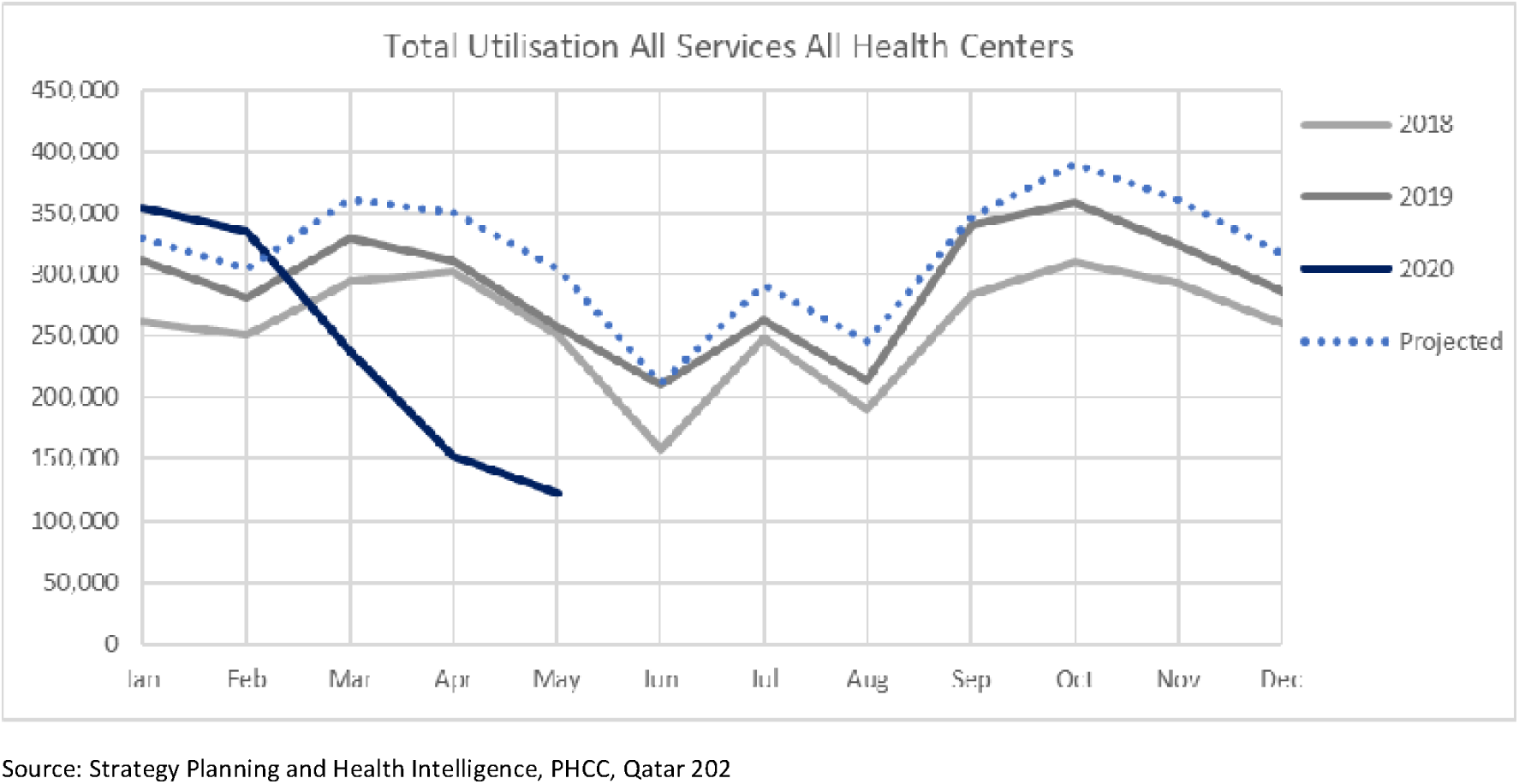
PHCC overall utilization for all services at all health centers between January 2018 and May 2020.

The overall utilization of all PHCC services across all health centers demonstrated a sharp decline during March to May 2020 in comparison to previous years and projected utilization. The reduction reached to 50% in in April 2020 in comparison to the previous two years (from 300,000 to 150,000 visits shown in figure No.4.

**Figure No.4:**
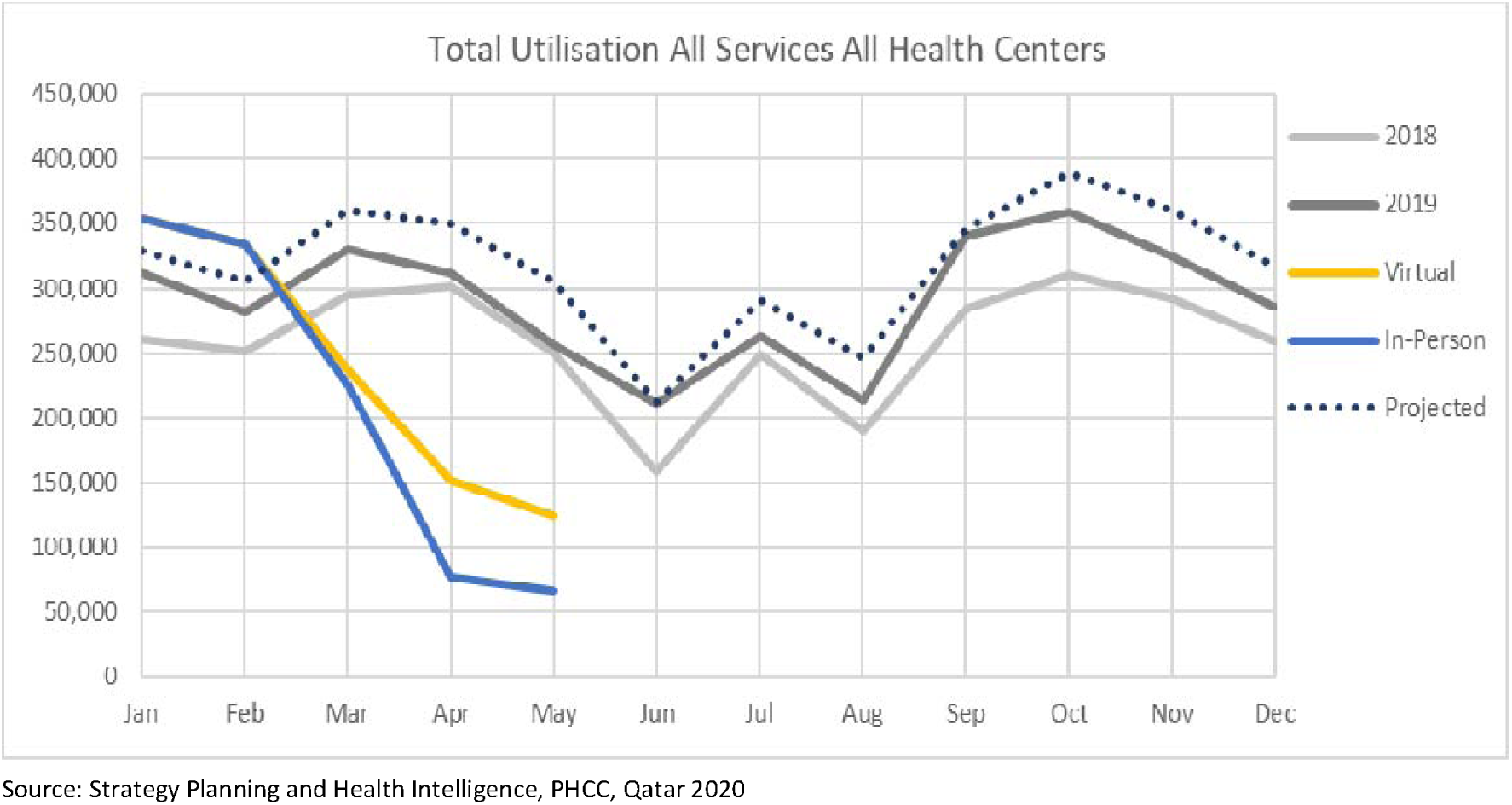
Total virtual consultation and in-person consultation utilization across PHCC health centers between January 2018 and May 2020.

### 3. PHCC alternative services provided due to COVID-19

On the 18^th^ of March 2020, PHCC commenced providing teleconsultation for all booked appointments with priority given for noncommunicable diseases (NCDs) patients. PHCC started as of 14^th^ of April 2020 to proactively calling all high-risk patients who didn’t have a booked appointment scheduled in 4 weeks’ timeframe. Priorities were given for patients aged 60+, NCD patients and pregnant women. Moreover, the PHCC established an inbound call centre on the 27^th^ of March 2020 to provide teleconsultation on the demand. The introduction of the virtual services had grown in utilisation volumes to the point where it made up 50% of the total April 2020 consultation volumes, and has been established to ensure urgent care is delivered appropriately where required however, most of the virtual consultation were categorised under general medicine as shown in figure No.4.

On the 24th of March 2020, PHCC started the implementation of home medication delivery service for the medication refills in collaboration with Qatar Post. The service targeted patients aged 60+ year, NCD and pregnant women to avoid the unnecessarily visits to the health centers (See figure No.5).

**Figure No.5:**
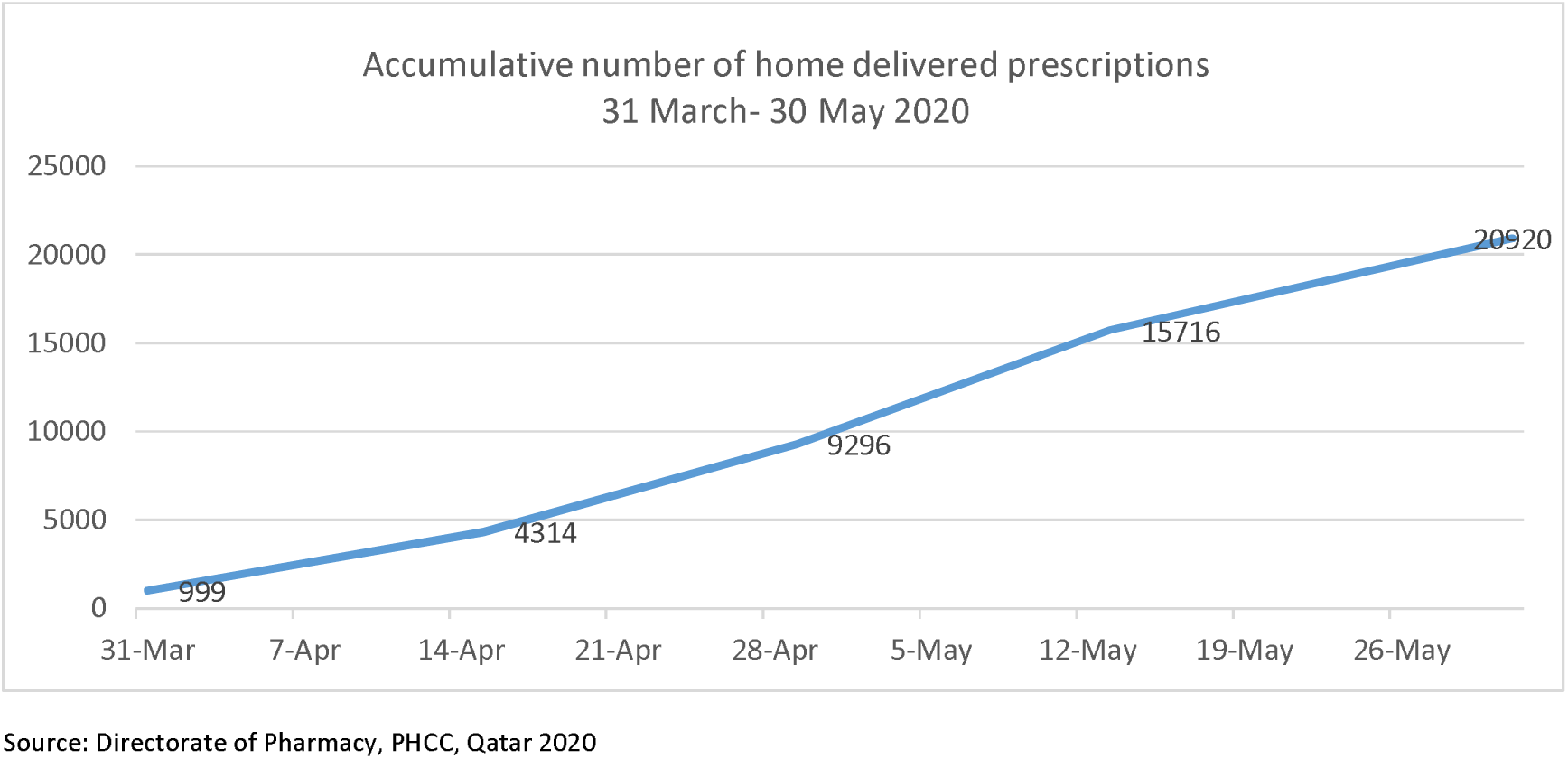
Home delivered prescriptions between 31st of March and 31st of May 2020.

## Conclusion & recommendations

The COVID-19 pandemic has impacted primary health care services in Qatar. This impact has both positive and negative sides for the services and patients. To decrease the risk of infection to the patients and health care workers, Primary Health Care Corporation (PHCC) in Qatar cancelled the appointments for some high-risk population e.g. NCD and antenatal visits, and the preventive visits e.g. screening and wellness. The latter reflected in a reduction of 50% in the overall utilization rate in comparison to the expected one for the month of April.

However, the PHCC has replaced the in-person consultations with telemedicine consultations. The pandemic affected the patients’ behaviour due to either adherence to the recommendations to stay at home, or their perceived risk of infection. Hence, the introduction of virtual services in March had an impact on the utilisation volumes to the point where it substituted 50% of the total April 2020 consultation volumes. Home medication refill delivery service for elderly, NCD patients and pregnant women was introduced at the end of March 2020. The service uptake reflects a steep incline in demand by the target population reaching up to 20,920 deliveries by end of May 2020. Although these services were included in the primary care plans, the national response to the pandemic had a positive impact on accelerating the implementation. Both home medication refill delivery and teleconsultation could play major role in the future to decrease the unneeded visits and make the primary care services more accessible for the community.

Meanwhile, PHCC continued to play an important role in screening for the COVID-19 suspected cases at the community through its COVID-19 centers and the remaining health centers with a total of 54824 swabbed conducted between the 14^th^ of March and the 15^th^ of June 2020. The latter represents almost 18% of the total number of tested people in Qatar. Additionally, PHCC through its testing sites managed to detect 13.4% of the total positive cases in Qatar.

These findings will pave the way for more research to understand in depth the effects of these findings on the behaviours and satisfaction from both the patients and primary care. In addition to understand the clinical outcomes on certain high-risk groups such as the people with chronic conditions.

## Data Availability

The volume data for the services utilization are available upon request

